# The effect of Health Literacy on COVID-19 Vaccine Hesitancy: The Moderating Role of Stress

**DOI:** 10.1101/2021.06.16.21258808

**Authors:** Huiqiao Zhang, Yue Li, Sihui Peng, Yue Jiang, Huihui Jin, Fan Zhang

## Abstract

**Background:** The COVID-19 vaccine is an essential means to establish group immunity and prevent the spread of the pandemic. However, the public’s hesitation has created major difficulties to the promotion of the vaccine. By investigating the relationship between health literacy and COVID-19 vaccine hesitancy, as well as the potential moderating role of stress, the present study would provide critical insights for tailoring vaccine-promotion strategies.

**Objective:** The two-fold research purpose is: i) address the effect of health literacy on people’s attitude toward COVID-19 vaccine, ii) clarify the role of stress in this effect.

**Methods:** With structured questionnaires, an online survey was conducted to evaluate general public’s COVID-19 vaccine hesitancy, health literacy, and perceived stress. In total, 560 responses were collected, and moderated regression analysis was conducted to test the effect of health literacy on vaccine hesitancy among people with different levels of stress.

**Results:** A total of 560 participants aged over 18 years were included in this study. About 39.8% of the respondents reported vaccine hesitancy, and this rate is higher among those aged 20-30 years old (83%) and female (71.3%). The results showed people with higher level of health literacy are less likely to have vaccine hesitancy (β =-2.00, 95%CI= [-3.00∼ -0.99]). However, this effect was only among those with lower to moderate level of stress (β =-3.43, p<0.001), among the people with high stress, no significant effect of health literacy was found (β =-0.53, p>0.05).

**Conclusions:** By focusing on the effect of health literacy on COVID-19 vaccine hesitancy, the findings showed education program increasing individual’s health literacy may also effectively reduce the public’s vaccine hesitancy and promote accepting attitude. However, for people with high level of stress, other health programs need to be developed to enhance their positive attitude toward the COVID-19 vaccine. In conclusion, promotion strategies should be tailored for different populations, with considering individual factors such as health literacy and stress.

## Introduction

Since March, 11, 2020, the World Health Organization (WHO) has declared COVID-19 as a pandemic, and up till 31 May, 2021, it has pointed to over 171 million infected cases and a million casualties in 222 countries and regions [1]. In addition to a public health crisis, COVID-19 also has immense impacts on global economy and financial markets due to reduced productivity, business closures, trade disruption and so on [2]. To mitigate the impact of this epidemic, building up the public’s immunity is a key preventive and preparatory measure. According to the past experiences of fighting against polio, smallpox, rabies, and other infectious diseases, vaccination was considered to be the most effective and economical approach to build up immunity [3]. However, group immunity requires at least 70% of the whole population to be vaccinated [4], and few countries have reached this criterion. In China, the vaccination uptake rate was only about 45.86% [5], and in France, it was 16.80%, in Brazil 10.57% and in South Africa it was 0.82% [6].Why is the public reluctant to take vaccine even when free supply is provided? A major obstacle is the high prevalence of vaccine hesitancy. Therefore, it is critical to understand the modifiable factors influencing individual’s vaccine hesitancy, which would provide critical insights for developing interventions to promote the public’s COVID-19 vaccine uptake.

Vaccine hesitancy, described as intentions to delay or refuse taking vaccination despite the availability of vaccination service [7]. Vaccine hesitancy is a concept that was affected by complex contextual influences, individual and group factors, and vaccine-specific characteristics [7].Compared with contextual influences and vaccine-specific characteristics, individual factors are more likely to be altered when developing health-promotion strategies. Individual factors include an individual’s knowledge, awareness, and motivation for disease prevention and health care, which are closely related with one’s health literacy. Health literacy (HL), defined as the cognitive and social skills that determine individual’s motivation and ability to gain access to, understand, and apply health information to promote and maintain health [8]. In the context of COVID-19, HL is operationalized as one’s health knowledge, awareness, and capacity to implement preventive measures such as vaccine uptake, hand hygiene, wearing face masks, or keeping social distancing. For example, previous studies showed that people with higher health literacy showed fewer unhealthy behaviors, i.e., reduced tobacco and alcohol use, as well as increased physical exercise, thus indirectly promoting health [9]. In particular, limited health literacy was also found to be one of the reasons why older adults are unwilling to take influenza vaccination [10], or other preventive measures such as wearing masks [11]. Recent studies on health literacy and the uptake of COVID-19 vaccine found that people with poor health literacy reported higher COVID-19 vaccine hesitancy [12]. Health literacy, particularly vaccine literacy, was related with Willingness to be vaccinated [13]. Geana et al., has also found that low health literacy is a related with the uncertainty or refusal to receive the COVID-19 vaccine [14]. However, Casigliani et al.,(2020) has found that health literacy was unrelated with the attitude towards COVID-19 vaccination, suggesting inconclusive relationship between health literacy and vaccine attitude [15]. The mixed findings could be driven by the potential moderators or mediators, for example, individual’s perceived stress. Previous study on healthcare workers showed that in the outburst of COVID-19 pandemic, nurses’ stress level could affect their intentions to take the vaccine. Factors such as lacking personal protective equipment [16], deployment to isolation wards [17], and the preventive policy of their organizations [18], could all contribute to intense stress among nurses. Furthermore, high stress was found to be associated with stronger COVID-19 vaccine uptake intention [19]. However, how stress interacts with individual factors such as health literacy in affecting the public’s vaccination attitude has remained unstudied.

The present study has a two-fold research purpose: 1) to investigate the effect of health literacy on people’s attitude toward taking the COVID-19 vaccine; 2) to address the interplay between stress and health literacy in this relationship. We hypothesized that 1) higher health literacy can effectively reduce COVID-19 vaccine hesitancy; 2) Stress plays a moderating role in health literacy and vaccine hesitancy. The findings would provide insights for developing intervention programs promoting the public’s vaccination acceptance and, hopefully, increasing the COVID-19 vaccine uptake rate and helping build up group immunity to effectively prevent the spread of COVID-19.

## Method

### Data collection

A cross-sectional survey was conducted to examine the relationships between health literacy, perceived stress and vaccine hesitancy among community population in mainland, China. With a data collection platform, Wenjuanxing (a survey service website similar to Qualtrics or SurveyMonkey, but tailored to Chinese users), an online anonymous survey was implemented from January-March, 2021. Five hundred and sixty-two responses were collected in total. With a screening criteria, two responses (0.4%) were removed due to the participant’s age was under 18 years old. Written consent form was obtained from the participants before they took part in the study. The survey was approved by the research ethical committee of Jinan University.

### Measurement

Participant’s demographic characteristics, including as age, sex, marital status, education level, chronic illness, and health behavior, were collected.

*COVID-19 vaccine hesitancy* was measured by a revised version of the Parents Attitudes About Childhood Vaccines (PACV[20]), It includes 7 items, for example, “Vaccine approved for marketing by an authority is safe”, and “You agree that vaccinate against COVID-19 is an important preventive measure”. With a 3-point Likert scale, responses ranged from “0” (agree or strongly agree), “1” (uncertain), and “2” (disagree or strongly disagree). The total score of vaccine hesitancy was obtained, which was transformed into an index with the formula [I = Mean *15]. After the transformation, the vaccine hesitancy index ranges from 0 to 30, with higher score indicating more vaccine hesitancy [20, 21]. The previous study conducted with Chinese sample has demonstrated acceptable validity, and good reliability [22]. A vaccine hesitancy index of 0 to 15 indicates “do not hesitate”, and 16 to 30 indicates “hesitant”. The Cronbach alpha in our sample is 0.637.

Health literacy was measured by a 12-item health literacy questionnaire (HL-SF12[23]), derived from the European Health Literacy Survey Questionnaire (HLS-EU-Q), with including 3 vaccine-related items. There were 15 items in total, including 4 items for each domain of health literacy, i.e., health care, disease prevention, health promotion, and 3 vaccine-related items (e.g., “I can understand why I need vaccination”). With a 4-point Likert scale, the responses ranged from “1” (very difficult) to “4” (very easy), and “do not know” was coded as missing value. The average score was obtained to indicate individual’s level of health literacy, with higher score suggesting better health literacy [24]. The Cronbach alpha in our sample was 0.936, suggesting good internal consistency.

Perceived stress was measured by the Chinese Perceived Stress Scales (CPSS). Previous research suggested it has good reliability and validity in community sample [25-27]. The scale has 14 items in total, such as “Feel unable to control important things in life”, “Feel that the problem continues to accumulate and it is difficult to solve”. With a 4-point Likert scale, the responses ranged from “1” (never) to “4” (frequently), and higher score indicates greater perceived stress.

### Statistical analysis

Data analysis was conducted by SPSS 21.0 and Process plug-in. A liner regression model was performed to assess the main effect of health literacy on vaccine hesitancy and the interaction with perceived stress. The results of both unadjusted and adjusted methods were displayed, and in the adjusted model, sex, education, age, marital status, chronic conditions, and health behavior were controlled.

## Results

A total of 560 valid responses (99.6%) were included in the analysis. The majority of the responders were female (70.0%), and the average age is 30.25 years (SD=13.92). Of the sample, 22.7% were married and the majority having university education or above (79.5%). The average vaccine hesitancy score was 12.11(SD = 5.69), with 39.8% participants hesitant. The HL mean score was 3.22 (SD = 0.46), suggesting most people choose between “easy” and “very easy” (The descriptive results was presented in Table 1)

**Table 1.** Descriptive statistics for all variables. Means and standard deviations reported for interval and ordinal variables, proportions for nominal or binary variables (N=560).

| Group | N (% of sample)<br>Mean ± S.D. | Range |
| --- | --- | --- |
| **Sex** |  |  |
| Male (0) | 168(30.0%) |  |
| Female (1) | 392(70.0%) |  |
| **Age** | 30.25 ± 13.92 | 18 - 90 |
| **Marital status** |  |  |
| Married (0) | 126(22.5%) |  |
| Single/divorced/separated/widowed/others (1) | 434(77.5%) |  |
| **Educational level** |  | 1 - 4 |
| Uneducated | 4(0.7%) |  |
| Primary school | 10(1.8%) |  |
| High school | 101(18.0%) |  |
| University or college and above | 445(79.5%) |  |
| **Chronic illness** |  |  |
| Yes (0) | 120(21.4%) |  |
| No (1) | 440(78.6%) |  |
| **Health behavior** |  |  |
| Frequent physical exercise | 200(35.7%) |  |
| Non-smoking | 520(92.9%) |  |
| Non-drinking | 517(92.3%) |  |
| Regular physical examination | 181(32.3%) |  |
| **Health Literacy** | 3.22 ± 0.46 | 1, 4 |
| **Perceived Stress** | 23.26 ± 7.87 | 0, 45.5 |
| **Vaccine Hesitancy** | 12.11 ± 5.69 | 0, 30 |

Table 2 presents the results of unadjusted and adjusted regression model predicting vaccine hesitancy. The results showed that both sex and age, were significantly associated with vaccine hesitancy. In particular, older age and being female are related with higher level of vaccine hesitancy. In the adjusted model, higher level of HL was associated with lower vaccine hesitancy (β = -2.00, 95%CI [-3.00∼ -0.99]), after controlling for sex, age, marital status, educational level, chronic condition, and the number of health behavior.

**Table 2.**
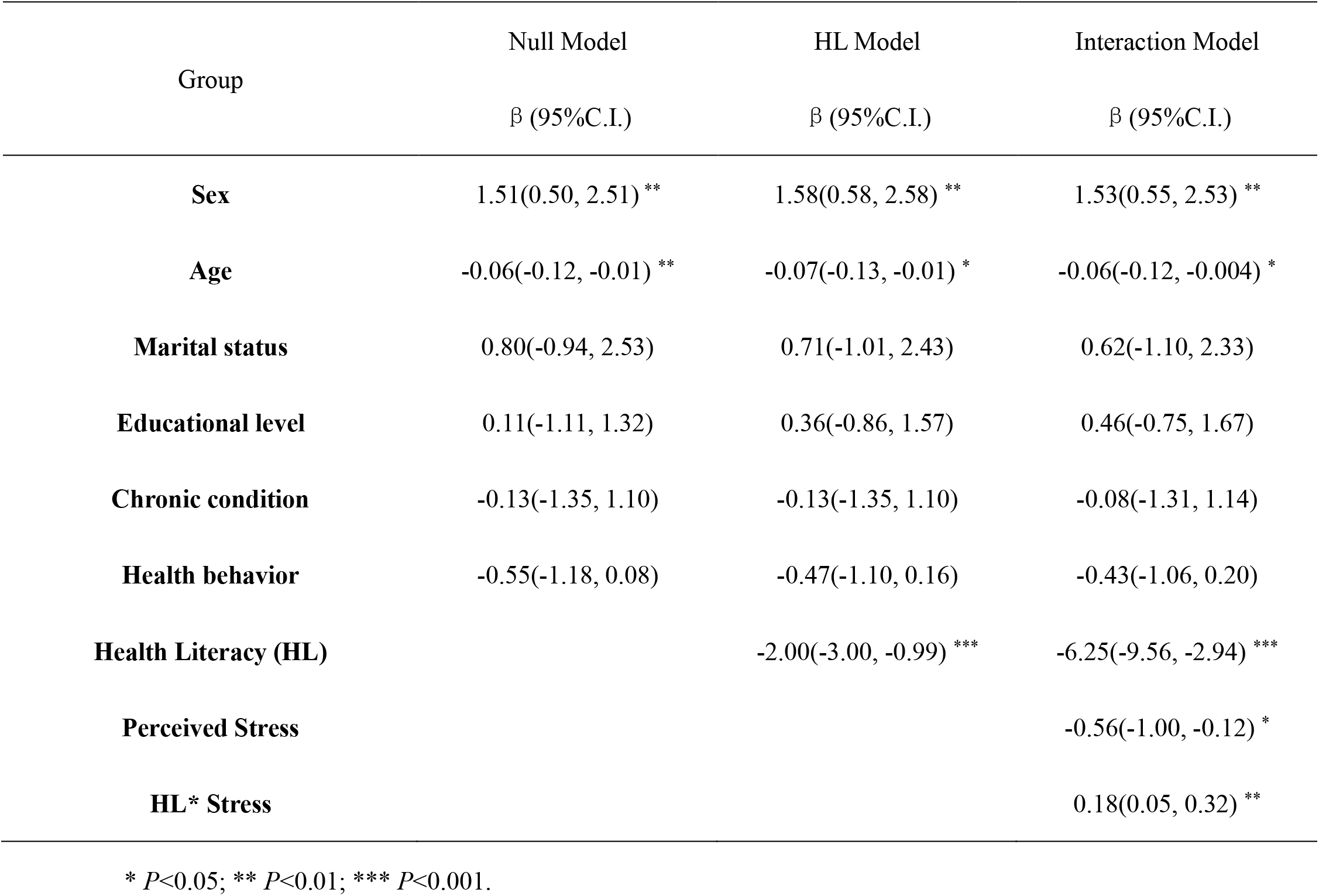
Results of unadjusted and adjusted model predicting vaccine hesitancy.

The Moderating model was constructed for health literacy, perceived stress, and vaccine hesitancy. The main effect of perceived stress was also significant (β = -0.56, 95%CI [-1.00, -0.12], suggesting that greater perceived stress was associated with lower vaccine hesitancy. The main effect was qualified by an interaction effect was found between HL and perceived stress, β_HL* Stress_ = 0.18 (95%CI [0.05∼0.32]), indicating the moderating role of perceived stress, and the effect of health literacy on vaccine hesitancy varies across different level of perceived stress. By probing the interaction effect, it was found that only when the perceived stress level was moderate or low, HL was associated with lower vaccine hesitancy (β =-3.43, *p*<0.001), and when the perceived stress level was high, the effect of HL became insignificant (β =-0.53, *p*>0.05).

**Figure 1.**
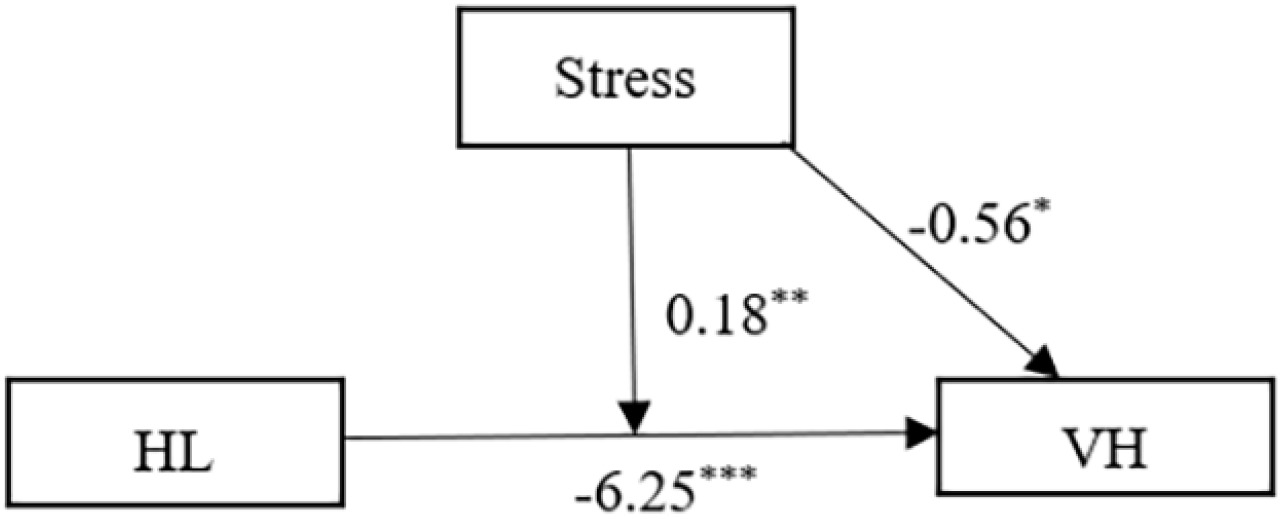
The moderation model of perceived stress, health literacy and vaccine hesitancy. * *P*<0.05; ** *P*<0.01; *** *P*<0.001

## Discussion

With a cross-sectional online survey, the present study has investigated how health literacy influences people’s COVID-19 vaccine hesitancy, and clarified the moderating role of individual’s stress level. The findings showed that, 39.8% of people reported COVID-19 vaccine hesitancy in our sample. People with higher level of health literacy are less likely to show hesitancy toward taking the COVID-19 vaccine. This effect was moderated by stress, such that only among people with low or moderate level of stress, health literacy was found to mitigate vaccine hesitancy.

Existing literature found that the rate of COVID-19 vaccine hesitancy varies across different countries and regions. With a sample of 2058 participants in China, the vaccine hesitation rate was 8.7% [28], which was much lower than that in our sample. However, the rate of vaccine hesitancy reported in the U.S.(42.4% [29]), in Turkey(44.5% [30]), and in Malta (48.2% [31]), were relatively close to that in our sample. In addition to the methodological inconsistence that may lead to different vaccine hesitancy rate, it also highly depends on the contextual factors. For example, when the infection threat was salient, people may report lower level of hesitancy. Otherwise, people are more likely to have concerns about the safety, adverse effect, and effectiveness of the vaccine. In our survey, we also asked about the reason people have for vaccine hesitancy, it showed that 62.5% were worried about the safety of vaccine, 46.4% had concerned that the COVID-19 vaccine is ineffective, and 14.3% were hesitant because of the inconvenience of taking vaccine.

Our results also showed that age, gender, chronic diseases are important factors predicting vaccine hesitancy. People aged from 20-30 seem to have the highest level of hesitancy while people aged above 60 showed the lowest. This is consistent with the findings in Malta [31], the US [32], UK and Ireland [33], that younger people tend to be more hesitant than older people. To have a better understanding on the age difference, we have conducted a focus group with 8 older adults aged above 65, and found that they showed higher trust on the authority, thus being more willing to comply the public policy regarding COVID-19, such as taking vaccine. Gender difference was also found on the attitude toward COVID-19 vaccine, that women showed more hesitancy than men in vaccination. In line with previous research [34-36], women tend to have greater concerns on the effectiveness of the COVID-19 vaccine, and their fear of injection could also result in vaccine hesitancy [37]. Moreover, our results showed that 32.5% of the respondents with chronic diseases were uncertain or unwilling to take a COVID-19 vaccine. Consistent findings in Italy also reported that patients with chronic diseases showed more concerns regarding the vaccine’s side effects, which reduced their willingness to take a vaccine [38]. Therefore, policy-makers should take age, gender, and individual’s health status into account, and tailor intervention programs for different populations.

The most important finding of the present study is that health literacy predicted lower level of vaccine hesitancy. On the one hand, health literacy helped individuals understand and evaluate the mechanism and effectiveness of COVID-19 vaccine, which enhance their self-efficacy in making decision of vaccination [39]. On the other hand, people with lower health literacy may be more susceptible to the false information about COVID-19 and vaccine in mass media, and find it hard to make vaccination decision. It may also fuel the mistrust for the authority and their advocacy of COVID-19 vaccine.

In addition, our finding showed that perceived stress was also associated with lower vaccine hesitancy and when the perceived stress is high, the effect of health literacy on reducing VH was not significant. According to the health belief model, higher stress may increase the perceived severity of and susceptibility to infection, thus increasing people’s willingness to take vaccine [40].In other words, both health literacy and stress could strengthen individual’s intention to take COVID-19 vaccine. The finding is consistent with the previous study that people with higher stress showed less vaccine hesitancy [19]. Although individuals with good health literacy were better able to make appropriate health decisions and perceive less hesitancy in taking vaccine [41], when perceived stress is high, it can trigger greater anxiety about the infection risk and interfere the decision-making process, thus masking the effect of health literacy.

## Limitations

Although the present study has identified the role of health literacy and stress in the COVID-19 vaccine hesitancy, there are a few limitations to be acknowledged. First, as a cross-sectional survey, the study only included vaccine uptake intention instead of the actual rate, which may raise potential questions regarding the findings. Follow-up study tracking participant’s vaccine behavior should be conducted to better address the role of health literacy in public’s vaccine practice. Second, participants were recruited at an online platform, which may inevitably result in selection bias. Older adults who may feel the most difficulties in making vaccine uptake decision may not be included in the sample. Third, our sample size is too small to capture the whole picture of Chinese society. Large-scale survey and cross-cultural comparisons are warranted to better understand the vaccine hesitancy of COVID-19.

## Conclusion

Our findings have highlighted that the effect of health literacy on reducing the COVID-19 vaccine hesitancy, particularly when the stress level is low or moderate. Given that the general level of health literacy has remained low in China, effort should be made to promote the public’s health literacy. Cultivating individual’s capacity to obtain, evaluate, and apply health information is vital for people to take preventive measures such as vaccine to protect themselves [42]. In addition, tailor-made program should be designed for groups with different gender, age, and physical conditions. For people with high stress, the effect of health literacy on vaccine attitude may become trivial, and specific strategies involving action cues should be developed to promote vaccine uptake and achieve the goal of herd immunity.

## Data Availability

Data can be achieved via emailing to

## Abbreviations

HL: health literacy
VH: vaccine hesitancy
SPSS: Statistical Package for the Social Sciences
CPSS: Chinese Perceived Stress Scales
PACV: Parents Attitudes About Childhood Vaccines
HL-SF12: 12-item health literacy questionnaire
HLS-EU-Q: European Health Literacy Survey Questionnaire
CI: confidence level
the US: The United States of America
UK: The United Kingdom of Great Britain and Northern Ireland

